# Prognostic Nutritional Index and Post-Stroke Depression: Insights from a Cross-Sectional Analysis of NHANES 2005–2018 Data

**DOI:** 10.1101/2025.10.12.25337842

**Authors:** Xueshan Jian, Yuxuan Ye, Xiaowan Lin, Yixiang Huang, Zhiru Zhang, Yuanyuan Zhang, Xiaona Tang, Rucheng Huang

## Abstract

**Background:** The Prognostic Nutritional Index (PNI), a composite biomarker that integrates nutritional reserve (albumin) and immune competence (lymphocyte count), has demonstrated prognostic utility across diverse clinical settings. However, its association with post-stroke depression (PSD) remains insufficiently elucidated, particularly in nationally representative large-scale cohorts.

**Methods:** For this investigation, we obtained data from the National Health and Nutrition Examination Survey (NHANES) spanning 2005–2018, a nationally representative cross-sectional survey. PNI was computed as 10 × serum albumin (g/dL) + 5 × total lymphocyte count (×10^9^/L). PSD was ascertained when participants had a Patient Health Questionnaire-9 (PHQ-9) score ≥10. Participants were categorized into quartiles according to their PNI values. Multivariable logistic regression (via three sequential models) and restricted cubic spline (RCS) analysis were employed to characterize the association between PNI and PSD. Subgroup and interaction analyses investigated potential heterogeneity across demographic subgroups (e.g., age, sex) and clinical subgroups (e.g., BMI, comorbidities).

**Results:** Among 32,220 participants, 220 (0.7%) fulfilled diagnostic criteria for PSD. In the fully adjusted Model 3, continuous PNI exhibited an inverse association with the risk of PSD (OR = 0.95, 95% CI: 0.92–0.99, p = 0.009). Participants in the highest PNI quartile (Q4: 44.02–56.01) demonstrated a 55% reduction in PSD risk compared to those in the lowest quartile (Q1: 12.01–40.01; OR = 0.45, 95% CI: 0.27–0.76, p = 0.003). Restricted cubic spline analysis verified a linear relationship between PNI and PSD (p = 0.825 for non-linearity). Subgroup analysis demonstrated a significant interaction solely with the poverty-income ratio (PIR, p = 0.005), and no heterogeneity was observed in other strata (all p > 0.05).

**Conclusion:** PNI exhibits a significant inverse linear association with PSD risk, supporting its role as a clinically feasible marker for PSD risk stratification. The PNI–PIR interaction highlights the need to contextualize PNI within socioeconomic frameworks when interpreting its prognostic value in PSD.

## 1. Introduction

Post-stroke depression (PSD), a prevalent and debilitating neuropsychiatric complication, affects approximately one-third of stroke survivors(1, 2). As global stroke burden escalates, PSD incidence rises concomitantly over time(3), characterized by heightened morbidity, mortality, and disability. As the global incidence of stroke continues to rise(4)—for instance, U.S. Centers for Disease Control and Prevention data show a 7.8% increase in stroke prevalence from 2011 to 2022, with a 14.6% surge in adults aged 18–44 years (2020–2022)(5)—research into PSD’s epidemiological characteristics and pathogenesis has grown increasingly critical. Clinically, PSD patients often present with neurological deficits and are more prone to motor and cognitive impairments, which in turn reduce quality of life, elevate mortality risk, and increase the family caregiving burden(4, 6, 7). Therefore, early identification and intervention of PSD are vital for enhancing patient outcomes. Existing studies indicate that PSD is closely associated with multiple factors such as inflammatory responses, neurotransmitter abnormalities, and endocrine disorders, though its specific pathogenesis requires further exploration(8).

Against this backdrop, indicators that integrate systemic health status—particularly those reflecting nutritional and immune status—have gained growing attention for their potential to identify PSD risk(9, 10). The Prognostic Nutritional Index (PNI), initially introduced by Onodera in 1984, was crafted to evaluate preoperative nutritional and immune status in surgical patients, providing a quantifiable gauge of this nutritional-immune interaction(11). It is computed as: PNI = 10 × serum albumin (g/dL) + 5 × peripheral lymphocyte count (10^9^/L), integrating two key biomarkers: albumin (a liver-synthesized protein that reflects nutritional reserves, metabolic stability, and anti-inflammatory capacity(12); low levels indicate malnutrition, chronic inflammation, or impaired liver function); and peripheral lymphocytes (core mediators of adaptive immunity that regulate inflammatory responses, support tissue repair, and modulate neuro-immune crosstalk; lymphopenia often signals immune exhaustion or systemic stress(13, 14)). Together, these components make PNI a reliable indicator of the body’s ability to cope with physiological stress—whether from acute injuries like stroke, chronic inflammation, or nutritional deficiencies(15–17).

PNI has been widely validated across clinical settings, demonstrating prognostic value in cancers, cardiovascular diseases, and neurodegenerative disorders—lower PNI levels correlate with poorer recovery outcomes, higher complication rates, and increased mortality(18–20). Its strength lies in its simplicity: based on routine laboratory tests (albumin and lymphocyte counts), it serves as a practical and cost-effective tool for assessing systemic health. Given that nutritional deficiencies and immune dysregulation are increasingly recognized as contributors to PSD—through their effects on neuroinflammation, neurotransmitter synthesis, and neuroplasticity— and PNI directly reflects nutritional and immune status, PNI may act as a potential marker for identifying high-risk individuals for PSD. However, large-scale epidemiological data linking PNI to PSD remain scarce, and its utility in this context is yet to be fully explored.

To fill this research gap, the present study utilizes nationally representative data drawn from the National Health and Nutrition Examination Survey (NHANES) spanning 2005 to 2018 to explore the association between PNI and PSD among stroke survivors. By employing multivariate regression models complemented with subgroup analyses, the study seeks to clarify whether PNI is linked to PSD risk and verify its potential as a clinically applicable indicator. Our study seeks to provide epidemiological evidence for PSD risk stratification, nutritional interventions, and comprehensive post-stroke care strategies.

## 2. Methods and materials

### 2.1. Study population

This study received ethical review and approval from the National Center for Health Statistics (NCHS) Ethics Review Board, and participants or their legal representatives furnished written informed consent(21, 22). We conducted an analysis of data from 7 cycles of NHANES (2005–2018), initially including 70,190 participants. Participants with missing critical data were excluded, as follows: (1) Age < 20 years (n = 30,441), as the study focused on adult stroke survivors; (2) Missing data on stroke history (n = 60), to ensure inclusion of only stroke survivors; (3) Missing responses to the Patient Health Questionnaire-9 (PHQ-9, used for PSD assessment; n = 5,606); (4) Participants with missing laboratory data required for PNI calculation (serum albumin or peripheral lymphocyte count; n = 1,863) were also excluded. Eventually, 32,220 participants were included in the final analysis. The participant selection process is illustrated in **Figure 1**.

**Figure 1.**
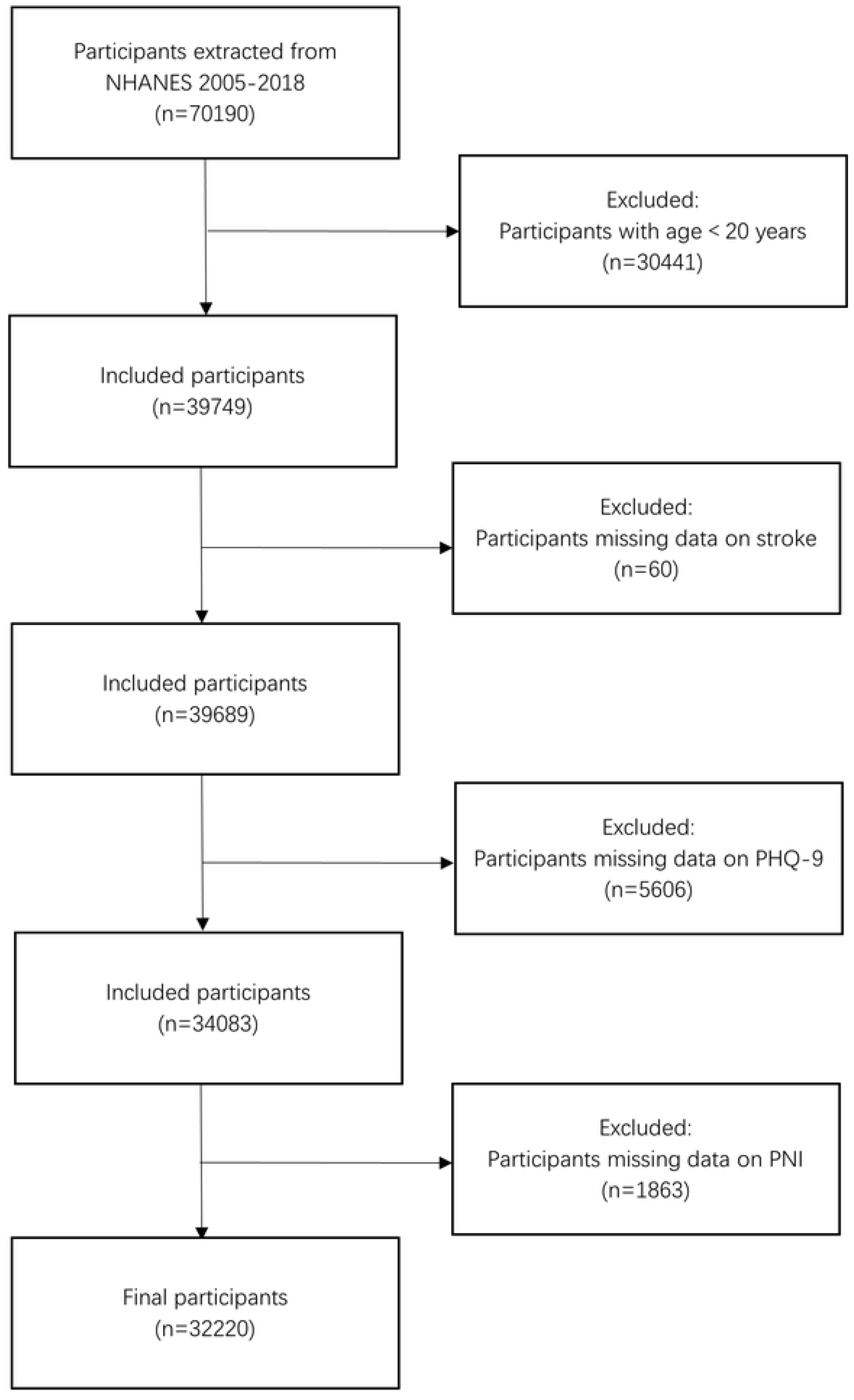
Flow diagram for sample selection from NHANES 2005-2018.

### 2.2. Assessment of PSD

Stroke history was determined based on participants’ self-reports in answer to an NHANES questionnaire item: “Has a doctor or other healthcare provider ever informed you that you had a stroke?” Depressive symptoms were assessed with the 9-item Patient Health Questionnaire (PHQ-9)—a tool developed to measure the frequency of depressive symptoms over the previous two weeks. Each of the nine items on the questionnaire is rated on a 4-point scale (0–3), where response choices correspond to: “not at all,” “several days,” “more than half the days,” and “nearly every day”—yielding a total score ranging from 0 to 27. In line with standard diagnostic criteria, a PHQ-9 score of ≥10 was employed to detect clinically significant depressive states. Participants were categorized as having PSD if they reported a history of stroke (self-reported) and a PHQ-9 score that met or exceeded the depressive symptom cutoff (≥10).

### 2.3. Assessment of PNI

The Prognostic Nutritional Index (PNI) serves as a composite indicator reflecting both nutritional and immune status. The formula to calculate PNI is as follows: PNI = 10 × serum albumin (g/dL) + 5 × peripheral blood lymphocyte count (10^9^/L). Peripheral blood lymphocyte counts were derived from the complete blood count (CBC) test. For NHANES data collection, automated hematology analyzers from Beckman Coulter were used—these instruments currently employ impedance and volume-based analytical methods. Further details are available in Chapter 7 of the NHANES Laboratory Manual. Serum albumin concentration—a well-established marker for protein nutritional status—was determined via standard biochemical techniques. More specifically, the bromocresol purple (BCP) dye-binding bichromatic endpoint assay was employed, which provides high levels of accuracy and reproducibility.

### 2.4. Covariate Selection and Definition

For the purpose of adjusting for potential confounding factors that could influence the association between PNI and PSD, we incorporated the covariates listed below: sex, age, race, education level, marital status, body mass index (BMI), poverty-income ratio (PIR), smoking status, hypertension, diabetes, congestive heart failure, and coronary heart disease. Age was stratified into three groups: <60 years, 60–74 years, and ≥75 years. Race was categorized into the following groups: Mexican American, non-Hispanic White, non-Hispanic Black, other Hispanic, or other races. Education level was classified into three categories: less than high school, high school or GED, or above high school. Marital status was categorized into three groups: married or living with a partner, widowed/divorced/separated, and never married. BMI— computed as weight in kilograms divided by the square of height in meters—was classified as normal (<25.0), overweight (25.0–30.0), or obese (>30.0). Socioeconomic status, assessed via PIR, was categorized as low (<1.3), moderate (1.3–3.5), or high (>3.5). Smoking status comprised three groups: current smokers (≥100 lifetime cigarettes and currently smoking), former smokers (≥100 lifetime cigarettes but not currently smoking), or never smokers (<100 lifetime cigarettes). Hypertension was defined if participants met any of the following criteria: (1) Average systolic blood pressure ≥140 mmHg; (2) average diastolic blood pressure ≥90 mmHg; (3) self-reported diagnosis of hypertension. Diabetes was diagnosed if participants met at least one of the following criteria: fasting plasma glucose ≥7.0 mmol/L, glycated hemoglobin (HbA1c) ≥6.5%, or self-reported physician diagnosis. Congestive heart failure and coronary heart disease were identified based on self-reported physician diagnosis. Detailed measurement protocols can be found in the NHANES technical documentation(http://www.cdc.gov/nchs/nhanes/).

### 2.5 Statistical analyses

Statistical analyses were conducted using IBM SPSS Statistics v26 and RStudio. Missing values in covariates were addressed through multiple imputation by chained equations (MICE), which involved 5 imputed datasets. The imputation model included all exposure (PNI), outcome (PSD), and covariate variables to preserve key multivariate relationships. Continuous variables were described as mean ± standard deviation (SD) for normally distributed data (normality evaluated through Shapiro-Wilk tests and visual inspection) or median (interquartile range, IQR) for skewed data; group comparisons were conducted with t-tests or Mann-Whitney U tests. Categorical variables were reported as frequencies (%) and analyzed using chi-squared tests. Multivariate logistic regression models were constructed to compute odds ratios (ORs) and 95% confidence intervals (CIs) for characterizing the PNI-PSD association. Three sequential models were specified: Model 1: Unadjusted (no adjustments). Model 2: Adjusted for sex, age, race, education level, and marital status. Model 3: Further adjusted for BMI, PIR, smoking status, hypertension, diabetes, congestive heart failure, and coronary heart disease. Restricted cubic spline (RCS) analysis was used to explore potential non-linear relationships between PNI and PSD risk, with adjustments for the same covariates as those in Model 3 to ensure consistency. Subgroup analyses, together with interaction tests (implemented via likelihood ratio tests for product terms in logistic regression models), were carried out to assess heterogeneity in the association across sex, age group, BMI, PIR, hypertension, and diabetes. Statistical significance was established at two-tailed p < 0.05 for primary analyses, and interaction p-values < 0.05 were considered to signify heterogeneous associations.

## Results

### 3.1. Baseline characteristics of NHANES participants (2005–2018)

Table 1 presents the baseline characteristics of NHANES participants stratified by PNI. A total of 32,220 participants were enrolled in this study, 220 of whom were identified as having post-stroke depression (PSD). The overall cohort had a mean age of (49.6 ± 17.8) years and a mean PNI of (42.1 ± 3.6). Participants were categorized into four PNI quartile groups: Q1 (12.01–40.01, n=8,060), Q2 (40.01–42.01, n=8,149), Q3 (42.01–44.02, n=7,958), and Q4 (44.02–56.01, n=8,053). PSD risk was significantly lower in Q2–Q4 than in Q1, with PSD prevalence declining gradually from 1.1% in Q1 to 0.2% in Q4. When compared with participants in the higher PNI groups (Q3–Q4), those in the lowest PNI group (Q1) were older (mean age: 53.0 ± 17.9 years vs. 44.0–49.5 years), predominantly female (67.2% vs. 32.8%–46.6%), and had lower educational attainment (proportion with education above high school: 50.2% vs. 52.7%–56.5%). This group was also more likely to be widowed/divorced/separated (28.0% vs. 14.5%–21.4%), have lower socioeconomic status (poverty rate [PIR<1.3]: 35.0% vs. 29.0%–29.9%), and higher body mass index (BMI) (prevalence of obesity [BMI>30 kg/m^2^]: 53.2% vs. 23.2%–33.9%). Regarding comorbidities, the lowest PNI group had a higher prevalence of hypertension (50.4% vs. 33.7%–41.4%), diabetes (25.4% vs. 11.6%–16.5%), congestive heart failure (5.9% vs. 1.6%–2.1%), and coronary heart disease (5.7% vs. 2.6%–3.7%) than the higher PNI groups. There were also significant differences in race distribution: the Q1 group had a greater proportion of non-Hispanic Black participants (29.2% vs. 13.8%– 17.2%), while it had a smaller proportion of non-Hispanic White participants (39.7% vs. 44.5%–47.5%). Smoking status differed significantly across groups (p=0.003) but showed no clear trend, as current smoking rates ranged from 19.3% to 21.1% across the four quartiles. All the aforementioned differences were statistically significant (p<0.001 unless otherwise noted).

**TABLE 1.**
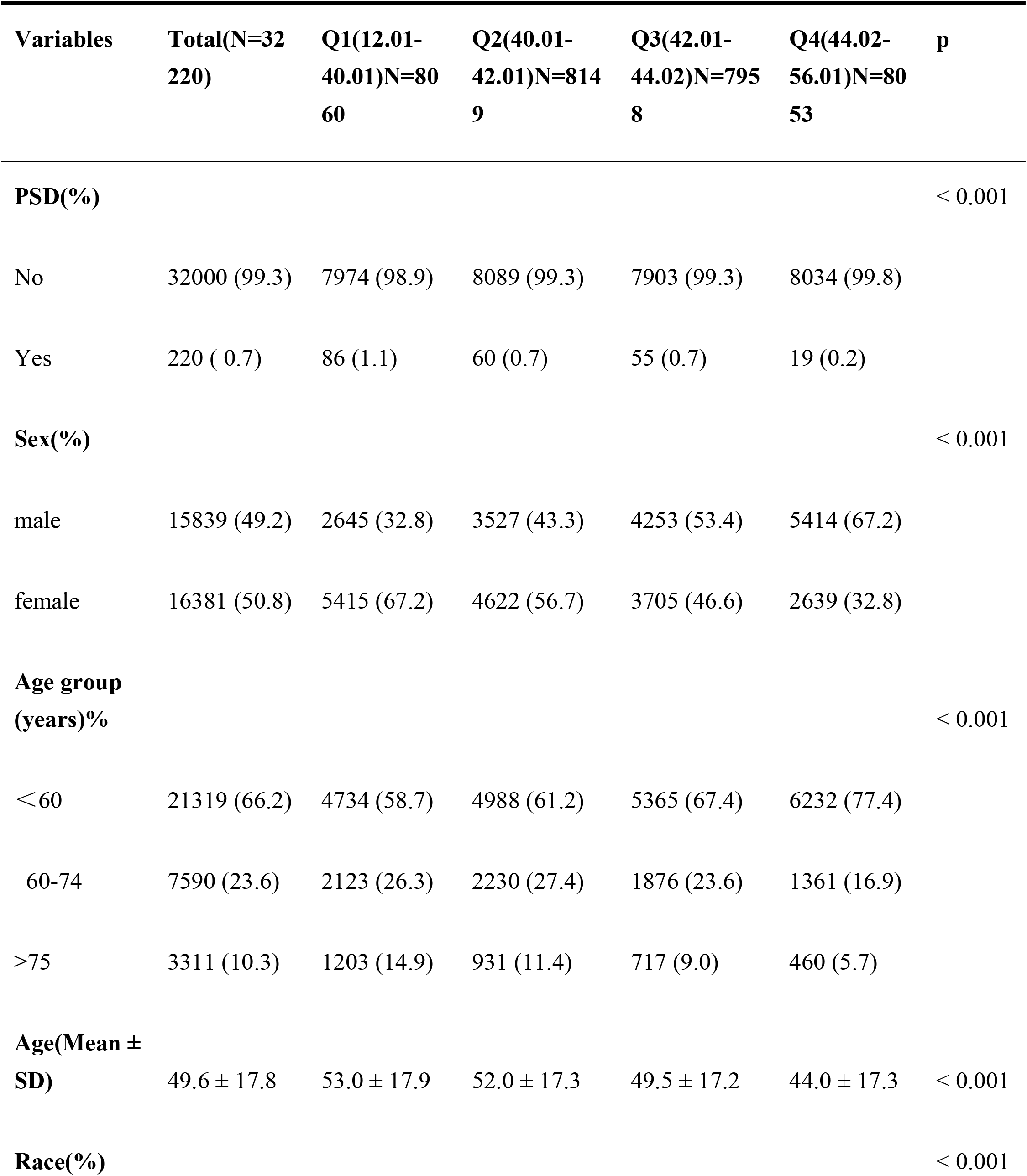
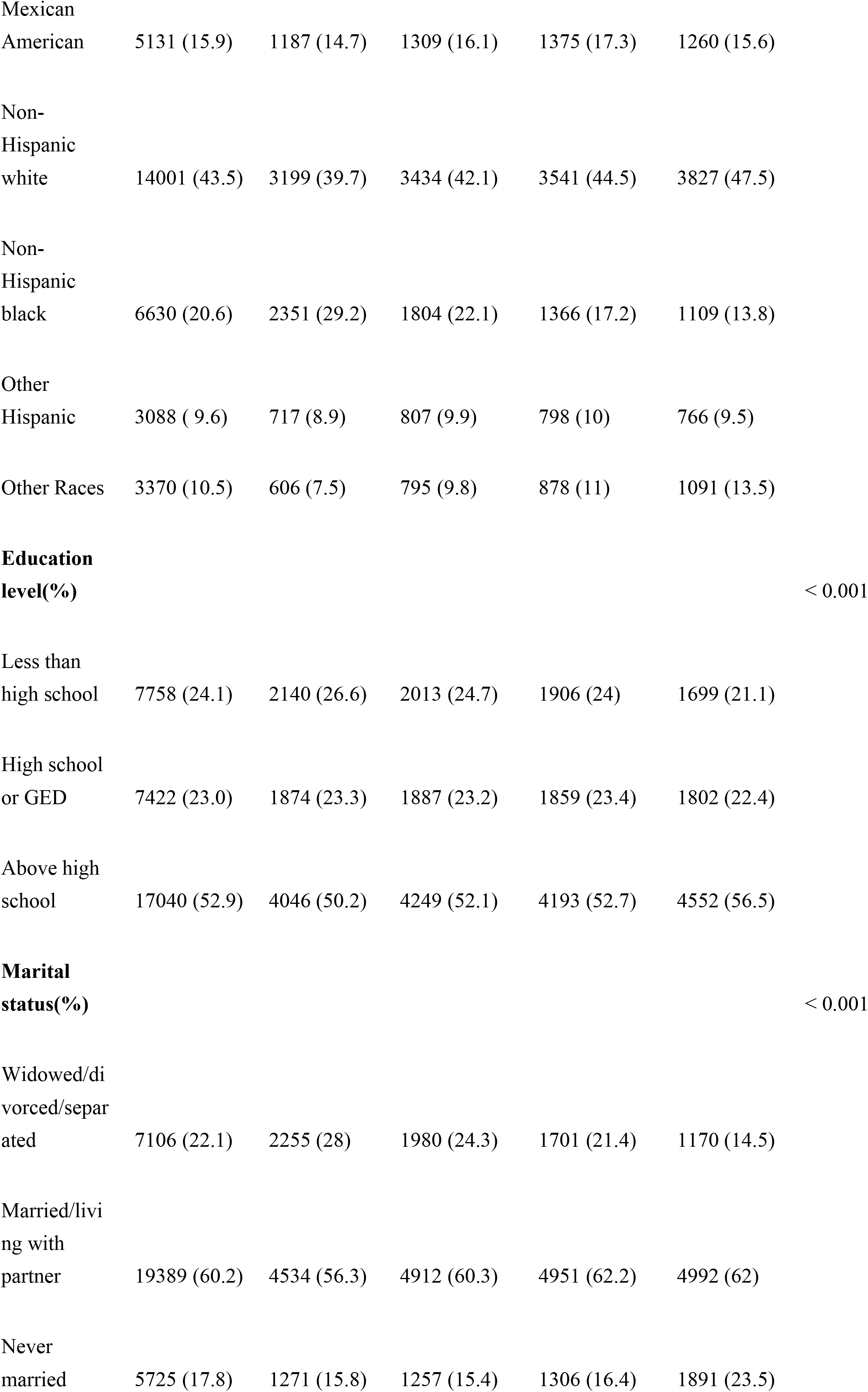
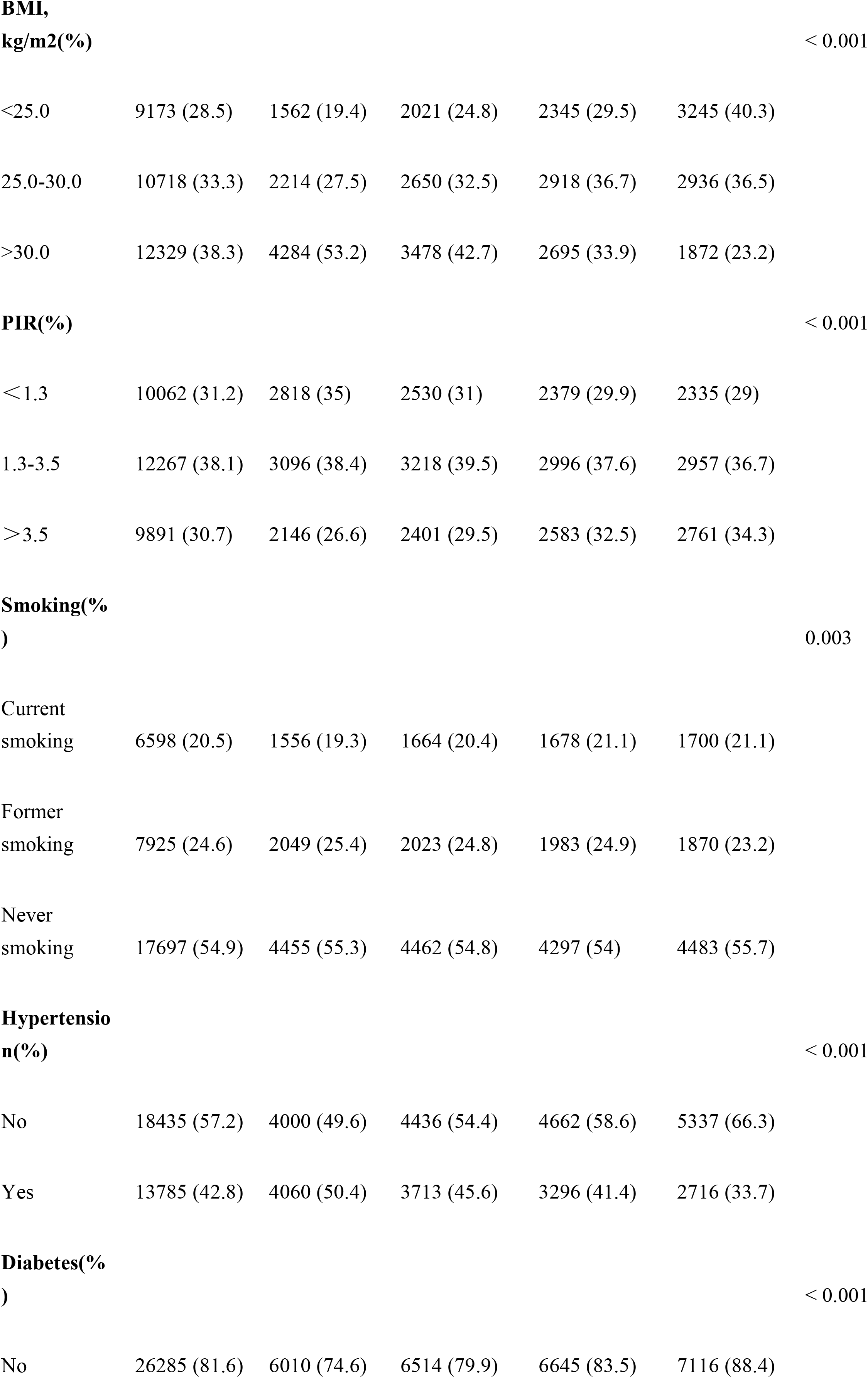
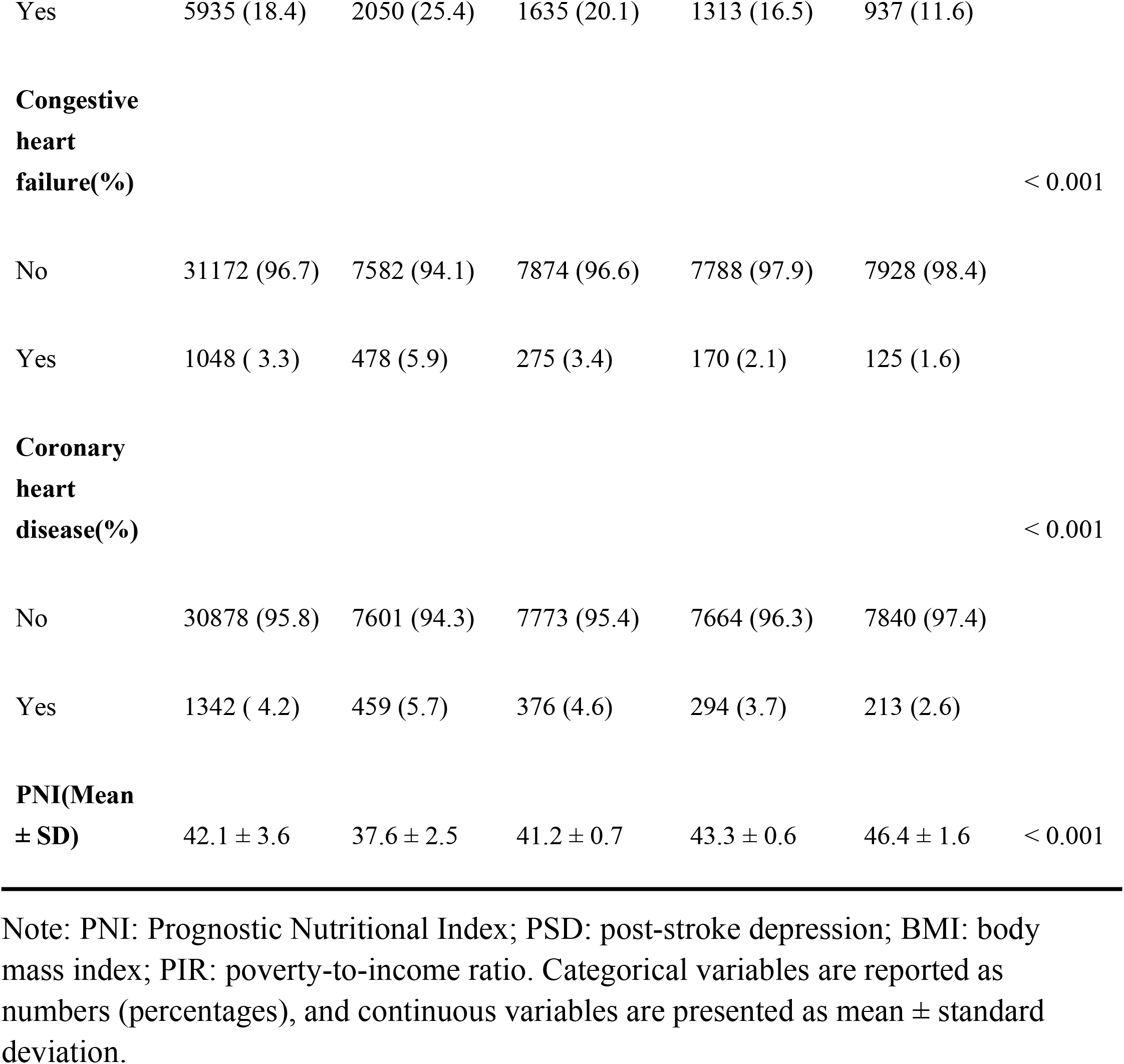
Characteristics of the study population from NHANES 2005–2018.

### 3.2 Association between PNI and PSD

Table 2 summarizes the findings from the multivariate logistic regression analysis exploring the relationship between PNI and PSD risk.

**TABLE 2.**
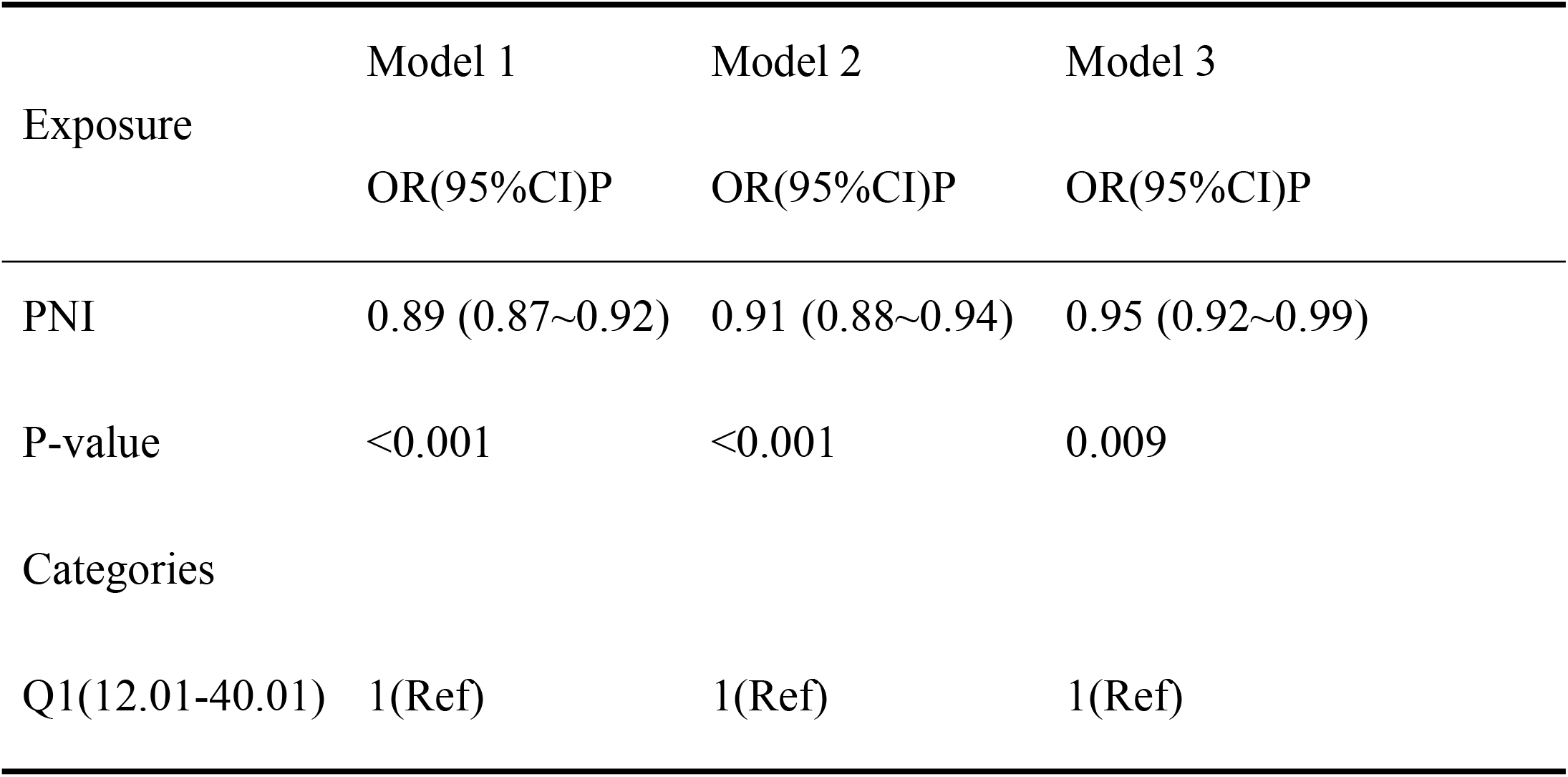
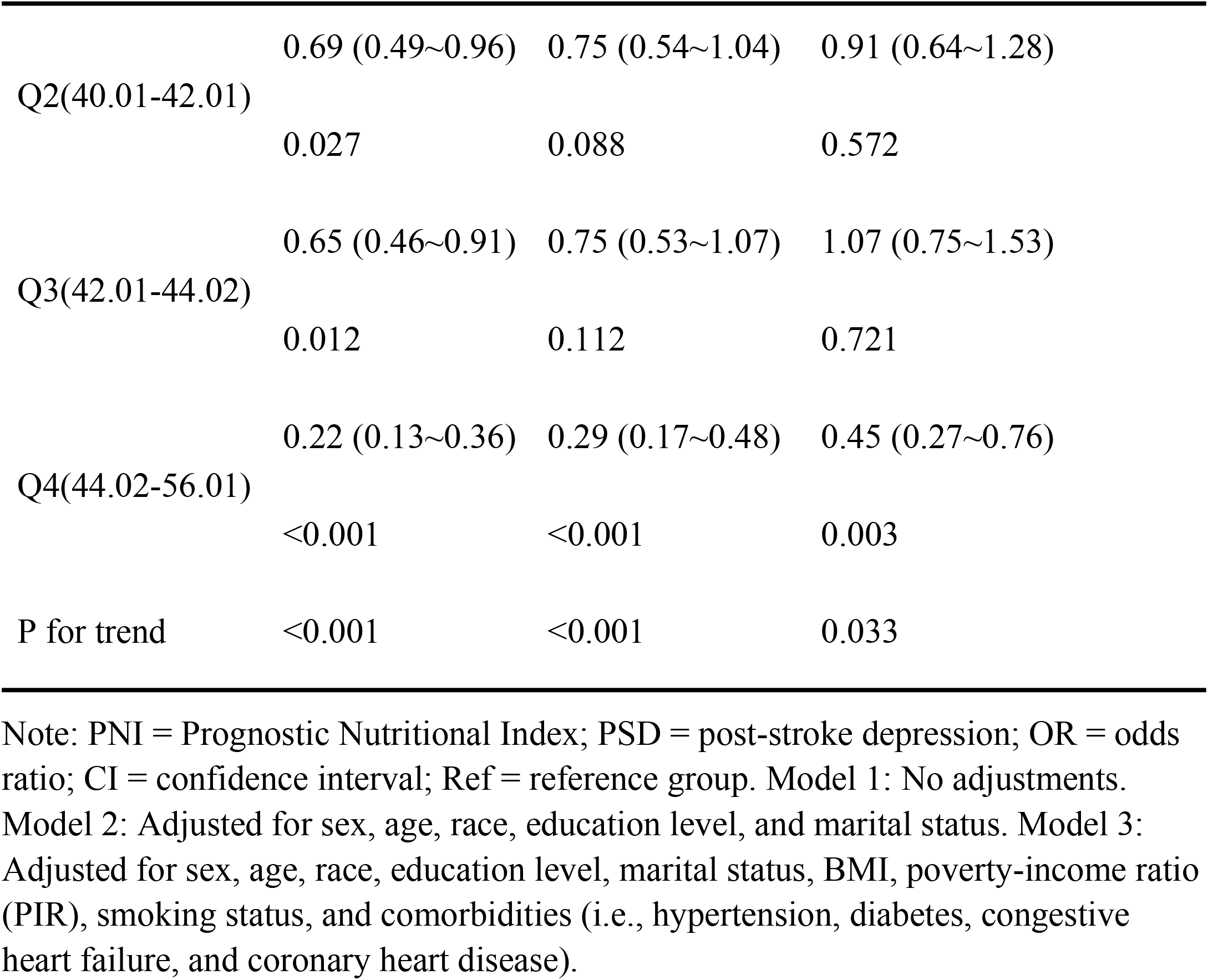
Logistic regression analysis of the association between PNI and risk of PSD.

Model 1 (Unadjusted): When treated as a continuous variable, PNI showed a significant negative correlation with PSD risk (OR = 0.89, 95% CI: 0.87–0.92, p<0.001). In the quartile-based analysis (with Q1 as the reference), PSD risk declined progressively across PNI quartiles: Q2 (OR = 0.69, 95% CI: 0.49–0.96, p=0.027); Q3 (OR = 0.65, 95% CI: 0.46–0.91, p=0.012); Q4 (OR = 0.22, 95% CI: 0.13–0.36, p<0.001). A trend test verified a strong inverse dose-response relationship (p for trend <0.001). Model 2 (Adjusted for Demographics): After adjusting for sex, age, race, education level, and marital status, continuous PNI maintained its protective effect (OR = 0.91, 95% CI: 0.88–0.94, p<0.001). Quartile-based analysis produced the following outcomes: Q2 (OR = 0.75, 95% CI: 0.54–1.04, p=0.088); Q3 (OR = 0.75, 95% CI: 0.53–1.07, p=0.112); Q4 (OR = 0.29, 95% CI: 0.17–0.48, p<0.001). The inverse trend remained highly significant (p for trend <0.001). Model 3 (Fully Adjusted): Further adjustment for BMI, PIR, smoking status, hypertension, diabetes, congestive heart failure, and coronary heart disease attenuated the association but retained statistical significance for continuous PNI (OR = 0.95, 95% CI: 0.92–0.99, p=0.009). Quartile-specific effects were as follows: Q2 (OR = 0.91, 95% CI: 0.64– 1.28, p=0.572); Q3 (OR = 1.07, 95% CI: 0.75–1.53, p=0.721); Q4 (OR = 0.45, 95% CI: 0.27–0.76, p=0.003). The persistent inverse trend (p for trend = 0.033) underscores that PNI acts as a robust independent protective factor against PSD.

### 3.3. Restricted cubic spline analysis

The results of the restricted cubic spline (RCS) analysis to examine potential nonlinearity in the PNI–PSD association are shown in **Figure 2**. RCS modeling was conducted to probe for such nonlinearity, adjusting for all covariates (consistent with Model 3). The nonlinearity test yielded a non-significant result (P=0.825), providing no statistical support for deviations from linearity. Visually, the spline curve (solid line) exhibited a consistent inverse trend across the central PNI range (30–45)—as indicated by the histogram overlay, which captured 76.3% of observations. Within this interval, the 95% confidence interval (shaded area surrounding the curve) remained narrow, reflecting robust precision in model estimation. Quantitatively, model-derived estimates confirmed a linear inverse association within this clinically relevant range: At the reference point (PNI=38), the estimated OR was 1.00 (95% CI: 0.92–1.08); At PNI=46, the OR declined to 0.82 (95% CI: 0.73–0.92), corresponding to an 18% reduction in PSD risk (95% CI: 8%–27%). These findings align with the linear association identified in logistic regression. At extreme PNI values (PNI<35, 3.8% of observations; PNI>50, 5.2%), the curve showed minor fluctuations, while the shaded 95% CI expanded sharply—reflecting imprecision from sparse data. Model outputs for these extremes (e.g., PNI=30: OR=1.30, 95% CI: 0.98–1.72; PNI=55: OR=1.12, 95% CI: 0.85–1.48) lacked statistical power to confirm meaningful risk changes. Collectively, the RCS analysis corroborates the primary finding: a continuous linear inverse association prevails across the clinically relevant PNI range (38–46), with no statistically validated nonlinearity or risk elevation at extremes.

**Figure 2.**
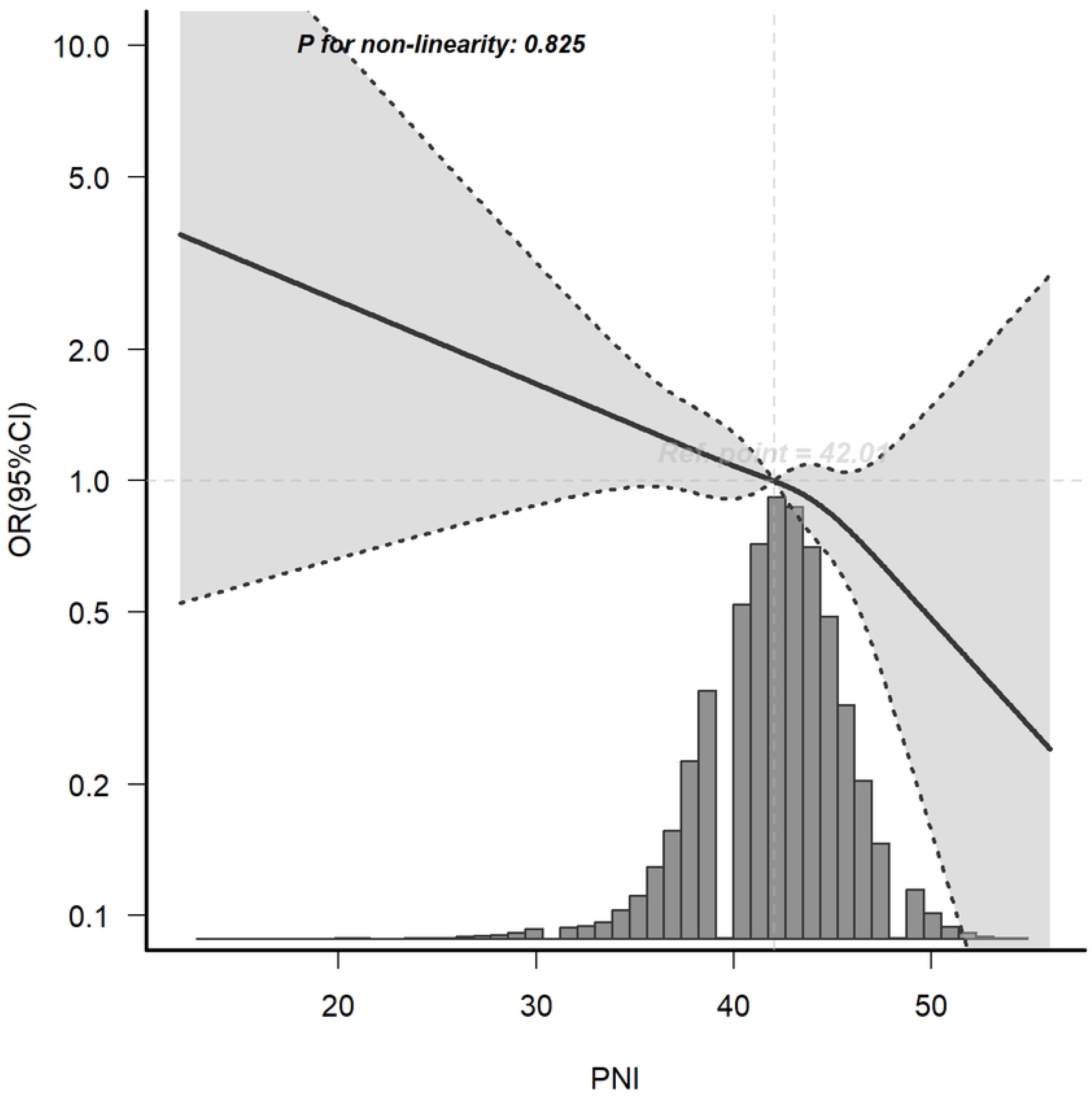
Association between PNI and PSD risk from restricted cubic spline (RCS) analysis. The solid line indicates estimated odds ratios (ORs), and the dashed lines represent 95% confidence intervals (CIs). Covariates for adjustment were consistent with those in Model 3 of the logistic regression analysis.

### 3.4. Subgroup and Interaction Analyses

Subgroup and interaction analyses exploring heterogeneity in the PNI–PSD association are presented in **Figure 3** (models adjusted for Model 3 covariates). Subgroup analysis identified significant negative PNI–PSD associations only in four subgroups: PIR > 3.5 (OR = 0.84, 95% CI: 0.76–0.93, p = 0.001); BMI > 30 kg/m^2^ (OR = 0.93, 95% CI: 0.89–0.98, p = 0.011); Diabetes (OR = 0.92, 95% CI: 0.87–0.98, p = 0.004); Hypertension (OR = 0.95, 95% CI: 0.91–0.99, p = 0.024). Interaction testing revealed a sole significant interaction between PNI and PIR (p = 0.005), with no interactions observed for sex, age, or other covariates (all p > 0.05).

**Figure 3.**
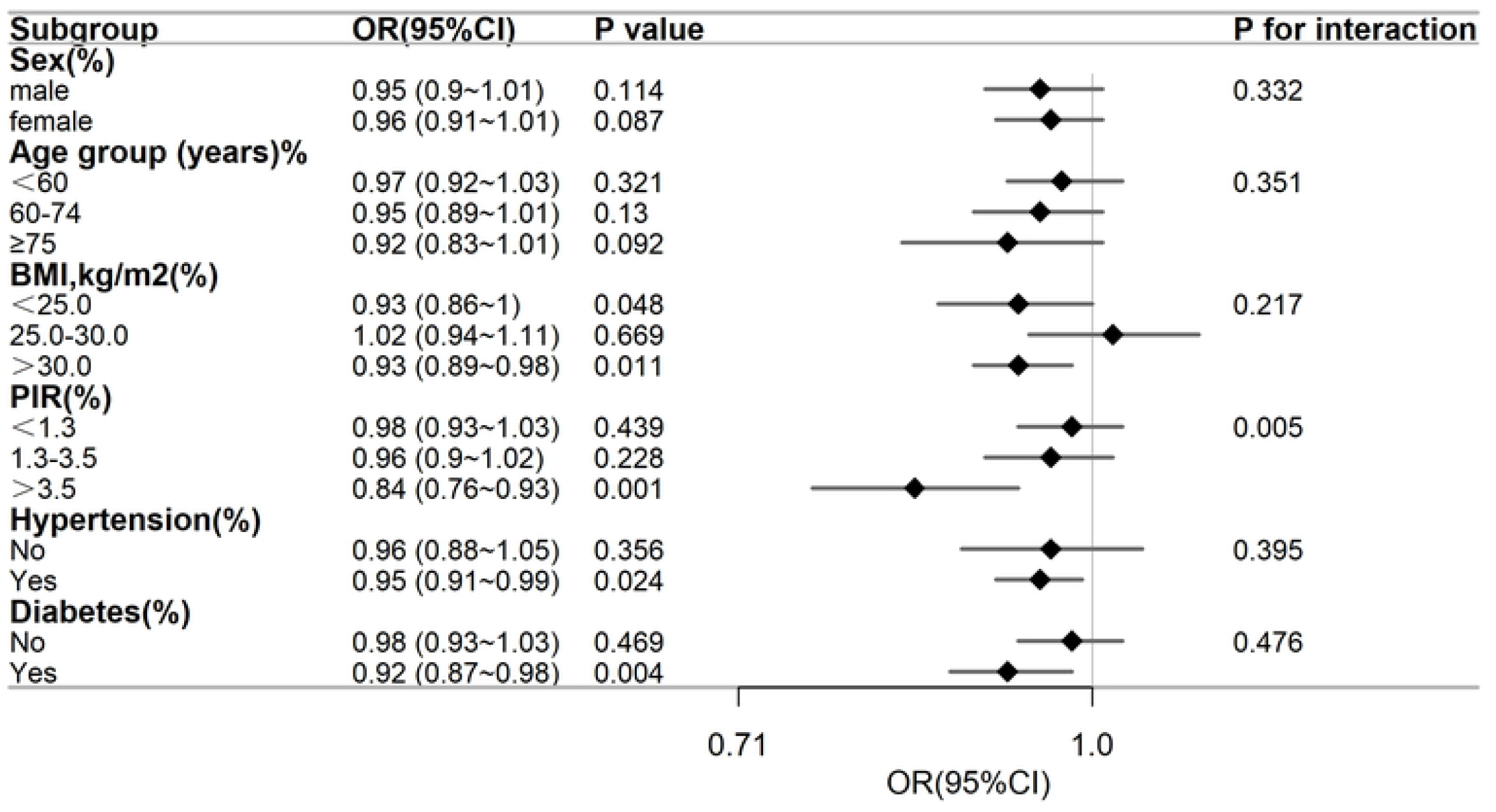
Subgroup analysis of the PNI–PSD association. Solid diamonds = odds ratios (ORs); horizontal lines = 95% confidence intervals (CIs). Significant associations (p < 0.05) are highlighted for PIR > 3.5, BMI > 30, diabetes, and hypertension, with a sole interaction for PIR (P = 0.005). Adjusted for Model 3 covariates.

## 4. Discussion

Our study identified an inverse association between PNI and PSD risk which stayed robust following strict adjustment for potential confounders. Notably, participants in the highest PNI quartile (Q4) exhibited a 55% reduction in PSD risk compared to those in the lowest quartile (Q1) in the fully adjusted model, underscoring the substantial protective effect of higher PNI. To our knowledge, this work represents one of the first large-scale epidemiological investigations leveraging the NHANES database to explore the PNI–PSD link. It aimed to address a critical research gap by systematically evaluating PNI’s prognostic value for PSD risk. Restricted cubic spline analysis, adjusting for all covariates, confirmed a linear PNI–PSD association (p for non-linearity = 0.825), with a consistent inverse trend across the clinically relevant PNI range (30–45, encompassing 76.3% of participants). This linearity bolsters PNI’s utility as a continuous biomarker for PSD risk stratification, whereas fluctuations at extreme PNI values (<35 or >50, 9.0% of observations) reflected sparse data, limiting interpretability. Subgroup analysis revealed the PNI–PSD inverse association was particularly pronounced in participants with PIR > 3.5, obesity (BMI > 30 kg/m^2^), diabetes, or hypertension (all p < 0.05). Interaction testing identified a sole significant interaction with PIR (p = 0.005), whereas no interactions emerged for sex, age, or other covariates—indicating the association’s stability across subgroups. The stronger protective effect in the high-PIR subgroup (PIR > 3.5) may involve plausible mechanisms: Biologically, higher socioeconomic status correlates with better nutritional intake and immune function, potentially enhancing PNI’s role in mitigating neuroinflammation and neurotransmitter dysregulation(23, 24); sociodemographically, greater healthcare access and lower exposure to chronic stressors may synergistically reinforce these effects(25, 26). Nevertheless, subgroup and interaction findings should be interpreted cautiously as preliminary, pending validation by well-designed prospective studies to confirm these relationships and dissect underlying mechanisms.

Plausible mechanisms underlying the PNI–PSD inverse association involve intertwined nutritional and immune pathways, as PNI integrates serum albumin (a cornerstone of nutritional status(27, 28)) and peripheral lymphocyte count (a proxy for immune competence(29)). Both components emerge as key modulators of PSD pathophysiology, offering mechanistic anchors for our findings(30, 31). First, serum albumin, a critical nutritional reserve, sustains neuroplasticity and dampens neurotoxicity(32–34). It preserves blood-brain barrier (BBB) integrity by limiting permeability to pro-inflammatory mediators(35–37)—a function pivotal post-stroke, where BBB disruption amplifies neuronal damage and depressive phenotypes(28, 38). Additionally, albumin facilitates tryptophan transport—the serotonin precursor central to mood regulation(39). Reduced albumin (as in low-PNI groups) may curtail tryptophan delivery to the brain, fostering serotonin depletion and elevated PSD risk(28). Animal models corroborate this, with albumin supplementation attenuating hippocampal inflammation and depressive-like behaviors in stroke paradigms(40).

Second, lymphocytes regulate neuroinflammation—a core driver of PSD pathogenesis. Post-stroke, pro-inflammatory (e.g., IL-6, TNF-α) and anti-inflammatory (e.g., IL-10) cytokine dysregulation disrupts mood-modulating neural circuits(41). Lymphocytes, particularly T cells, constrain this imbalance by suppressing excessive microglial activation(42)—a process compromised in individuals with lymphopenia (and thus low PNI). Clinical studies link post-stroke lymphopenia to heightened IL-6 levels and more severe depressive symptoms(43), aligning with our observation that lower PNI correlates with increased PSD risk. Notably, our RCS analysis confirmed linearity across the clinically relevant PNI range (30–45), reinforcing that even marginal PNI increments confer incremental PSD protection. This aligns with prior work showing nutritional optimization (e.g., protein supplementation) and inflammation modulation (e.g., addressing lymphopenia via anti-inflammatory strategies) reduce PSD risk(30, 44).

Despite these mechanistic insights, several caveats warrant caution: (1) NHANES assesses PSD via self-reported PHQ-9 scores, which—while validated for population-level screening—carry a risk of underestimating severe cases or missing subclinical presentations; (2) PNI quartiles in this study were sample-specific and did not align with clinical nutritional thresholds (e.g., PNI < 40 for moderate malnutrition). Given NHANES targets the general population (not clinical cohorts), PNI’s role in PSD should be interpreted with this stratification in mind; (3) PNI overlooks other nutritional facets (e.g., micronutrient levels like vitamin D, a neuroinflammation modulator(45, 46)) and immune cell subtypes (e.g., regulatory T cells, critical for neuroprotective immunity(47), limiting a holistic understanding of its role; (4) Stroke history was self-reported, which may cause misclassification bias, though NHANES self-reported stroke data have previously shown reasonable agreement with medical records; (5) Unmeasured confounders—such as post-stroke antidepressant use or stroke severity, which is a robust PSD predictor not captured in NHANES—may bias the observed association.

Future research should build on these findings by: (1) Conducting prospective cohort studies to establish causal temporality (i.e., whether PNI changes precede PSD onset); (2) Integrating PNI with complementary biomarkers (e.g., serum IL-6, hippocampal volume via imaging) to refine PSD risk stratification; (3) Exploring tailored nutritional-immune interventions (e.g., albumin-enriched diets, immune-modulating therapies) in low-PNI individuals to reduce PSD incidence.

## 5. Conclusion

Our nationally representative cross-sectional study demonstrates a significant inverse linear association between PNI and PSD risk, where the highest PNI quartile (Q4) confers a 55% reduction in risk compared to the lowest (Q1). Restricted cubic spline analysis corroborates linearity across the clinically relevant PNI range (30–45), with the inverse trend consistently observed across subgroups—notably stronger in individuals with high socioeconomic status (PIR > 3.5). PNI, derived from routine clinical tests, emerges as a clinically accessible indicator for PSD risk stratification. Future research should prioritize prospective cohorts to establish causal temporality and mechanistic studies to validate PNI-targeted interventions for PSD prevention.

## 7. Author Contributions

XJ: Conceptualization, Data curation, Formal analysis, Methodology, Writing – original draft, Writing – review & editing. YY: Conceptualization, Formal analysis, Methodology, Validation, Writing – original draft. XL: Data curation, Formal analysis, Methodology, Writing – original draft. YH: Data curation, Formal analysis, Methodology, Writing – original draft. ZZ and YZ: Conceptualization, Methodology, Writing – original draft. XT: Funding acquisition. RH: Conceptualization, Funding acquisition, Project administration, Supervision, Writing – original draft, Writing – review & editing. All authors have read, reviewed, and approved the final manuscript.

## 9. Acknowledgments

All authors wish to express sincere gratitude to the National Health and Nutrition Examination Survey (NHANES) for making open-access data available, which has been instrumental in supporting this research.

## 10. Data Availability Statement

The datasets analyzed in this study are publicly available from the National Health and Nutrition Examination Survey (NHANES) database [https://www.cdc.gov/nchs/nhanes/index.htm]. Specific datasets used in this analysis can be identified by the survey years and component modules as detailed in the manuscript’s methodology. Further inquiries regarding the derived datasets used in this study can be directed to the corresponding author.

## 11. Ethics statement

This study was carried out in line with the principles of the Declaration of Helsinki. All protocols related to the National Health and Nutrition Examination Survey (NHANES) received approval from the National Center for Health Statistics (NCHS) Research Ethics Review Board, and all participants gave written informed consent.

## 12. Generative AI statement

The authors confirm that no generative artificial intelligence tools were used in the production of this manuscript.

